# Spatial variation in delayed diagnosis of visceral leishmaniasis in Bihar, India

**DOI:** 10.1101/2024.03.22.24304732

**Authors:** Emily S Nightingale, Joy Bindroo, Pushkar Dubey, Khushbu Priyamvada, Aritra Das, Caryn Bern, Sridhar Srikantiah, Mary M Cameron, Tim C D Lucas, Graham F Medley, Oliver J Brady

**Affiliations:** Department of Infectious Disease Epidemiology, London School of Hygiene and Tropical Medicine, London, UK; Independent; formerly affiliated with Bihar Technical Support Program, CARE-India Solutions for Sustainable Development, Patna, India; Department of Epidemiology and Biostatistics, University of California, San Francisco, CA, United States; Department of Disease Control, London School of Hygiene and Tropical Medicine, London, UK; Department of Health Sciences, University of Leicester, Leicester, UK; Department of Global Health and Development, London School of Hygiene and Tropical Medicine, London, UK

**Author notes:** Corresponding author: Department of Infectious Disease Epidemiology, London School of Hygiene and Tropical Medicine, London, UK. E-mail address (E. S. Nightingale).

## Abstract

**Background:** Visceral leishmaniasis (VL) is a debilitating disease and without treatment, a fatal disease which burdens the most impoverished communities in northeastern India. Control and ultimately, elimination of VL depends heavily on prompt case detection. However, a proportion of VL cases remain undiagnosed many months after symptom onset. Delay to diagnosis increases the chance of onward transmission, and poses a risk of resurgence in populations with waning immunity. We checked the spatial variation of delayed diagnosis of VL in Bihar, India and aimed to understand the potential driving factors of delayed diagnosis.

**Methods:** The spatial distribution of diagnostic delays was explored using a Bayesian model fit to geo-located cases using the Integrated Nested Laplace Approximation (INLA) approach, assuming days of delay as Poisson-distributed and adjusting for individual-(age, sex, HIV) and local-level (recent incidence, vector control, health facility access) characteristics. Residual variance was modelled with an explicit spatial structure. Cumulative delays were estimated under different scenarios of active case detection coverage.

**Findings:** The 4,270 cases analysed were prone to excessive delays outside existing endemic “hot spots”, beyond the focus of interventions. Cases diagnosed within recently-affected blocks and villages experienced shorter delays on average (by 13% 95% Credible Interval [2.9% - 21.7%] and 7% [1.3% - 13.1%], respectively) than those in non-recently-affected areas.

**Interpretation:** Delays to VL diagnosis when incidence is low could influence whether transmission of the disease could be interrupted or resurges. Prioritising and narrowing surveillance to high-burden areas may increase the likelihood of excessive delays in diagnosis in peripheral areas. Active surveillance driven by observed incidence may lead to missing the risk posed by as-yet-undiagnosed cases in low-endemic areas, and such surveillance could be insufficient for achieving and sustaining elimination.

**Funding:** The Bill and Melinda Gates Foundation.

## Introduction

Control of visceral leishmaniasis (VL) on the Indian sub-continent depends on prompt detection and treatment of cases through recognition of clinical symptoms or screening in affected areas. Early symptoms of VL are non-specific (including fever, fatigue and weight loss) and, especially where VL awareness is low, misdiagnosis is common. As a result, those afflicted may go undetected for several months or in extreme cases, undetected for years, despite the presence of active detection measures. Evidence suggests that longer time to diagnosis is associated with increased mortality risk^1^ and undetected cases serve as reservoirs of infection in their community, allowing transmission to persist and forming a barrier to achieving and sustaining elimination ^2^.

An improved programme of active case detection (ACD) ^3^ was initiated during 2016-2017 and its efficacy in reducing overall time to diagnosis has been demonstrated. However, a non-negligible proportion of cases are diagnosed several months after onset of symptoms. A study by Dubey and colleagues ^4^ reported that during the first 19 months of improved ACD in the Indian state of Bihar, 66% of diagnosed cases reported onset of symptoms greater than 30 days prior to diagnosis, and 10.5% greater than 90 days prior. Le Rutte and colleagues ^5^ estimated in 2017 that elimination of VL in the Indian sub-continent could be achieved by 2020 with sufficient coverage of vector control, “*provided that the average onset-to-treatment (OT) time does not exceed 40 days”.* The persistence of this minority of cases with long delays to diagnosis therefore deserves further investigation.

Barriers to diagnosis of VL have been previously investigated in several studies. VL burden is broadly associated with the most socially and economically disadvantaged communities in India ^6^ and, despite government compensation for expenditure to access VL diagnosis and treatment, patient costs remain an important barrier ^7^. Mondal and colleagues ^8^ screened households in villages sampled from endemic districts in Bangladesh, India and Nepal, finding a high proportion of undiagnosed cases in districts not well-served by health care facilities and a lower proportion in districts with greater availability of VL care (i.e. districts considered affected/endemic in which the elimination programme is active).

Rahman and colleagues ^9^ interviewed VL patients in Bangladesh and found logistical barriers to prompt diagnosis such as remoteness of the health centre, wet season transport limitations, restricted ability to travel due to and limited availability of RK39 rapid diagnostic tests (for VL diagnosis) in the area. This was combined with lack of understanding due to illiteracy, lack of recent incidence and preference for first consulting more local traditional healers. In Bihar there is also widespread use of private and informal health practitioners which can cause additional delays ^10^.

The same study by Dubey and colleagues ^4^ explored patient characteristics associated with longer delays between symptom onset and diagnosis among all cases of VL diagnosed between January 2018 and June 2019. It was concluded that younger age and detection via active surveillance were associated with shorter delays, while male sex and HIV positivity were associated with longer delays.

What has not been considered in previous literature is where geographically individuals are experiencing excessive delays, in relation to each other and in relation to the activities of the control programme. Control and surveillance of VL in Bihar is targeted according to recently observed incidence ^11^, resulting in a spatially-varying intensity of intervention. This work aims to investigate the spatial distribution of delays in diagnosis and understand some of its potential driving factors in the State of Bihar, India.

## Methods

### Data sources

This work is based on secondary analysis of data collated for a previous study ^4^ evaluating active case detection measures. Case reports of individuals diagnosed between Jan 01, 2018 and July 31, 2019 (N = 5,030) were cross-referenced with suspect case registers over the same period in order to identify the route of detection for each patient as active (via targeted surveillance) or passive (self-referral).

Low specificity of the recommended rapid diagnostic test (RK39) requires that only cases suffering at least 14 days of fever are suspected for VL and eligible for testing ^3^. The primary outcome was therefore defined as the reported duration of fever prior to diagnosis beyond the standard criteria of 14 days, hereafter referred to as “diagnosis delay”. This was considered theoretically avoidable delay within the current guidelines. Cases diagnosed within 14 days of fever onset were not considered comparable to the rest of the population and excluded.

### Village locations

Location data are available for every village with at least one case reported to the Kala-Azar Management Information System (KAMIS) from 2013-18. Cases described above were linked to their resident village and corresponding GPS location via a unique ID. Location data were predominantly unavailable for villages not affected between 2013-18.

### Health facility access

Capacity for diagnosis and treatment of VL is not consistent across all health facilities in Bihar, as treatment centres were originally established to be near the most affected villages ^12^ (*Supplementary figure S1 (A))*. A tool developed by The Malaria Atlas Project (MAP) was used to estimate minimal travel time between villages and the available diagnosis and treatment facilities by relative “accessibility” ^13^, accounting for distance and ease of travel (*Supplementary figure S1 (B)*).

### Baseline model structure

Reported diagnosis delay (in days) for each case, *Y_i_*, is assumed to be Poisson-distributed with mean λ*_i_*, with independent and identically distributed observation-level random effects (OLRE) to account for overdispersion ^14^. The model is fitted within a Bayesian framework using the Integrated Nested Laplace Approximation (INLA) approach.

Formally,

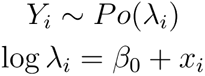

Where

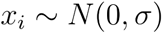

with a penalised-complexity hyperprior (20) set on the standard deviation σ, such that *P*[σ > 1] = 0.01. This penalises deviation from the simplest case in which the standard deviation is equal to 0 (i.e. constant) and specifies that the variance of these random effects is not expected to be greater than 1.

It is common for self-reported duration data to exhibit “heaping”, in which individuals show a preference for certain (usually rounded) intervals of time, and there has been suggestion that this behaviour may bias parameter estimates ^15^. The final model was therefore refitted with a binary outcome of delay exceeding 30 days, to assess the robustness of inferred covariate effects.

### Covariates

Covariates at both individual and village level were considered within three domains: patient, village risk awareness and village accessibility. Characteristics of the patient included age (standardised), sex (male / female), HIV status (positive / negative at diagnosis), marginalised caste status (scheduled caste or tribe / other), previous treatment for VL or post-kala azar dermal leishmaniasis (PKDL) (yes / no), occupation (none / unskilled / skilled / self-employed or salaried) and route of detection (ACD or passive/self-reported). Village characteristics were defined under two domains. Block endemicity (endemic / non-endemic), targeting of indoor-residual spraying (IRS) (yes / no) and village incidence of VL (non-zero / zero) in the previous year (2017) were considered indicators of ‘risk awareness’ in the local population. Estimated travel time (minutes) to the nearest diagnostic or treatment facility and diagnosis during the rainy season (June - September) were defined under the domain of ‘accessibility’. Both ACD and IRS are incidence-targeted interventions, triggered by incidence during the last three years. As such, these variables are expected to be to some extent correlated with 2017 village incidence. Estimated covariate effects are presented as risk ratios (RRs) with 95% credible intervals (CrI).

### Variable selection

The association of each covariate with observed delay was explored through univariately within the baseline model structure. Multivariate models were then fit for each domain in turn, and significant covariates selected based on the adjusted coefficients’ 95% CrI. A full model was then fit to include the selected covariates in all three domains.

### Spatial analysis

The correlation between delays experienced in nearby villages was modelled with a spatially-structured random field over the GPS locations for all villages, using the INLA-SPDE approach for estimation ^16,17^. This approach approximates a spatially-continuous field via stochastic partial differential equations (SPDE) across a triangular mesh. A prior structure which penalises complexity was also assumed for the hyperparameters of this component (range and standard deviation). A range of prior specifications for the SPDE model were explored to assess sensitivity to this choice and are illustrated in *Supplementary figure S7*.

A spatial field was initially added to the baseline, OLRE-only model, to characterise the spatial pattern in the absence of the explanatory power of the covariates. Each covariate domain was then reintroduced in turn and finally in combination, resulting in the following structure:

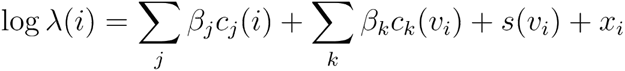

Where *c_j_*(*i*) are individual-level covariate values for case *i, c_k_*(*v_i_*) are village level covariate values for the village *v_i_* of case *i*, *s*(*v_i_*) is the spatial random field and *x_i_* the OLRE.

The contribution of each domain of covariates in explaining the spatial pattern of delays was explored via the percentage change in mean absolute value (MAV) across the fitted spatial field when each covariate domain was reintroduced. The percentage change in MAV of the OLRE was also calculated to assess the contribution of each in explaining the non-spatially-structured residual variation.

### Model assessment

The value of including both covariates and an explicit spatial structure was assessed via Widely Applicable (also known as Watanabe-Akaike) Information Criterion (WAIC) and leave-one-out (LOO) cross-validation, relative to the baseline OLRE-only model. Model predictions were compared on the logarithmic score (logs) ^18^ and on the Brier score ^19^ for classification of delays greater than 30 days. Spatial and non-spatial cross-validation approaches were compared to assess the contribution of the spatial random field to prediction (see *Supplementary Materials B)*.

#### Final model prediction

The expected extent of excessive delays from the selected model were mapped over all affected districts. Predictions were calculated for a fine grid of points across the area, reflecting the expected delay for an arbitrary individual at that location, otherwise comparable on all covariates. The posterior distribution is summarised by a mean and an exceedance probability with a threshold of 30 days and plotted to form a smooth map. In particular, regions in which the predicted exceedance probability is above 0.5 (i.e. where delay longer than 30 days is more probable than delay within 30 days) are highlighted.

#### Impact of ACD

To explore the potential impact of extending or restricting ACD across endemic and non-endemic regions of Bihar, hypothetical delays were predicted under two scenarios of ACD coverage among the individuals in this study (0% and 100%). Predicted days of delay where either no or all cases were detected via ACD were compared to the expected delays with ACD as originally observed. The difference in terms of total person-days of delay was stratified by the endemicity of the block and summarised over 10,000 posterior samples to capture uncertainty.

#### Ethical statement

Ethical approval was obtained from the London School of Hygiene and Tropical Medicine ethics committee for this study (ref:26841) and from the National Vector Borne Disease Control Programme in India (NVBDCP) for the work of the broader SPEAK India research consortium. The ethics committee of the All India Institute of Medical Sciences-Patna approved the ACD effectiveness evaluation protocol for analysis of data from the Kala-Azar Management Information System (KAMIS); no new data were collected under the research protocol.

#### Statistical analysis

All analyses were performed in R version 4.1.2 (2021-11-01). The written code has been made available at https://github.com/esnightingale/vl-spatial-diagnosis-delay.

## Results

### Data cleaning

Of 5030 patients diagnosed with VL between Jan 01, 2018, and July 31, 2019, 649 residents of villages with no known GPS location and one with an assumed erroneous GPS location substantially (>10km) beyond the state boundary were excluded. Two patients were removed from KAMIS due to recognition of an error therefore were also excluded. A further 84 were excluded due to missing HIV status, caste status, occupation or VL/PKDL treatment history. HIV status had the greatest proportion of missingness at 1.3%. Excluding incomplete observations had negligible impact on the distribution of delays, with equal means (31 days) and quartile ranges (11-44 days) before and after exclusion (*Supplementary table S1*).

Twenty four cases (0.5%) reported fever duration less than 14 days. Overall, patients diagnosed with less than 14 days of fever were younger, less likely to be female, more likely to reside in VL-endemic blocks and closer in travel time to diagnostic and treatment facilities (*Supplementary table S2*).

A summary of the data cleaning process is illustrated in *Supplementary figure S2* and a comparison of included and excluded cases presented in Supplementary tables *S1* and *S2*.

### Descriptive

4,270 VL patients diagnosed within the study period and with complete covariate information and linked to a GPS-located village were included for analysis. These had reported duration of fever ranging from 14 to 510 days at the point of diagnosis. The geographic spread and distribution of diagnosis delay for included patients is illustrated in *Figure 1B*.

**Figure 1:**
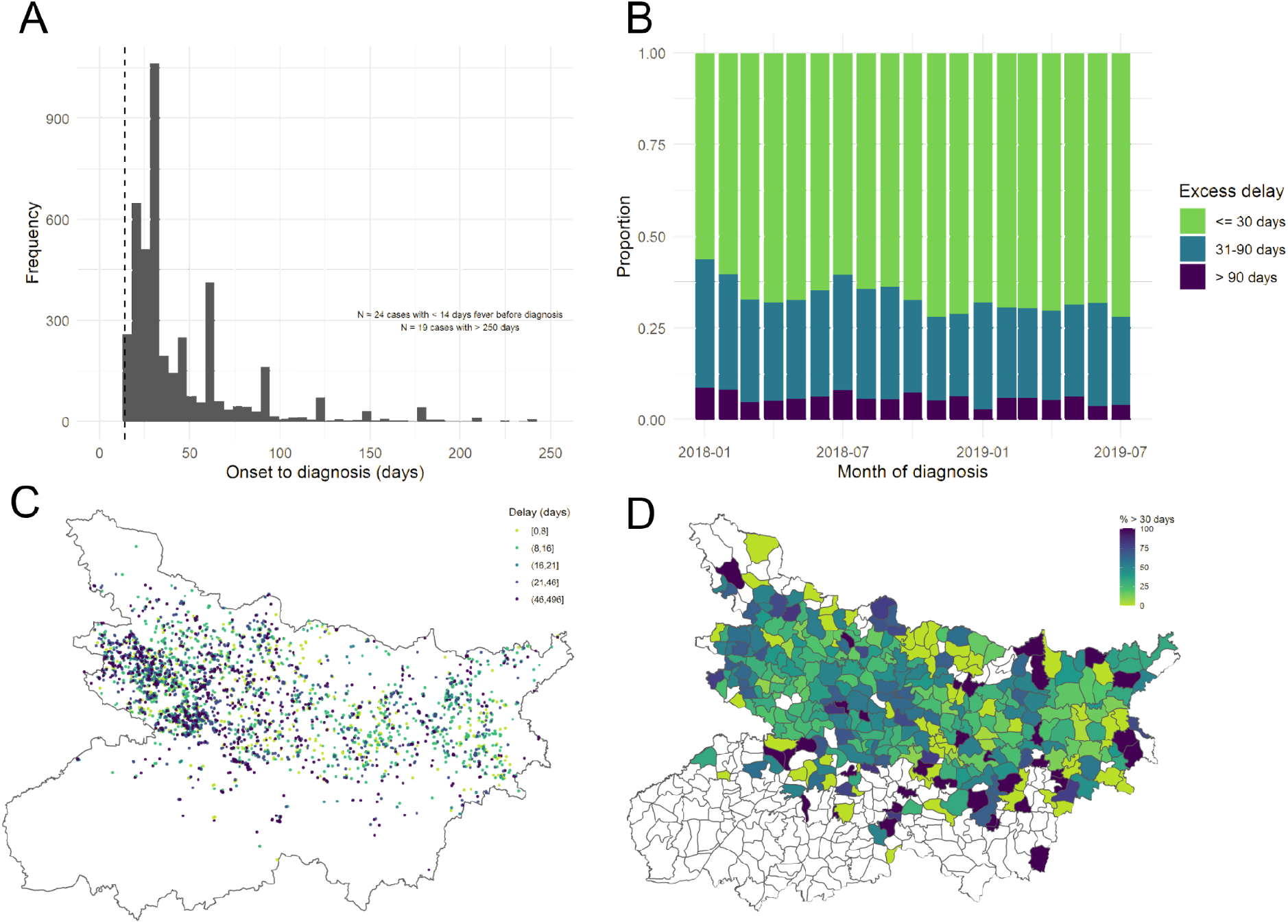
**(A)** Distribution of reported days from onset of fever to diagnosis for all initially included cases. The dashed line marks the 14 day criteria for diagnosis. Note the visible heaping in reported duration, indicating a preference for 30 day intervals. Panel **(B)** illustrates how the proportion of cases experiencing excessive delays varies by month of diagnosis. Panels **(C)** and **(D)** illustrate the geographic distribution of reported diagnosis delays, according to GPS location of resident village (after exclusion of < 14 day durations) and by resident block.

A descriptive summary of characteristics of included patients is presented in Table 1, and an illustration of the full correlation matrix between all considered covariates is shown in Supplementary figure S3.

**Table 1:** Descriptive summary of characteristics of 4,270 VL patients included in the analysis.

| Variable |  | N | Delay, mean (SD) | Delay > 30 days, N (%) |
| --- | --- | --- | --- | --- |
| Sex | Female | 1829 | 30 (34.4) | 623 (34) |
|  | Male | 2441 | 31.9 (41.2) | 814 (33) |
| Age (years) | < 13 years | 1154 | 25.9 (30.4) | 327 (28) |
|  | 13-25 years | 1056 | 27 (32) | 303 (29) |
|  | 26-42 years | 1031 | 35.4 (43.1) | 392 (38) |
|  | > 42 years | 1029 | 36.6 (45.8) | 415 (40) |
| Scheduled caste or tribe | No | 2778 | 32.1 (40.7) | 958 (34) |
|  | Yes | 1492 | 29.2 (33.9) | 479 (32) |
| Occupation | Unemployed | 2506 | 29.9 (35.8) | 818 (33) |
|  | Unskilled | 1213 | 32 (41.1) | 421 (35) |
|  | Skilled | 272 | 33.5 (37.4) | 99 (36) |
|  | Self-employed/salaried | 279 | 34.7 (48.8) | 99 (35) |
| HIV status | Negative | 4112 | 30.1 (36.1) | 1356 (33) |
|  | Positive | 158 | 55.1 (73.7) | 81 (51) |
| Previous VL/PKDL treatment | No | 3896 | 30.8 (37.3) | 1308 (34) |
|  | Yes | 374 | 34.2 (48.7) | 129 (34) |
| Detection route | Passive (self-report) | 2557 | 35.2 (41.6) | 1004 (39) |
|  | Active | 1713 | 24.8 (32.3) | 433 (25) |
| Block endemic in 2017 | No | 2332 | 34.4 (43.8) | 847 (36) |
|  | Yes | 1938 | 27 (30.4) | 590 (30) |
| Village IRS targeted in 2017 | No | 993 | 33.8 (42.1) | 359 (36) |
|  | Yes | 3277 | 30.2 (37.3) | 1078 (33) |
| Village incidence > 0 in 2017 | No | 1929 | 34.5 (42.9) | 726 (38) |
|  | Yes | 2341 | 28.2 (34.2) | 711 (30) |
| Travel time to nearest diagnosis facility | < 15 minutes | 2662 | 31.1 (37.3) | 910 (34) |
|  | 15-30 minutes | 1299 | 31.2 (41.6) | 421 (32) |
|  | > 30 minutes | 309 | 30.7 (34) | 106 (34) |
| Travel time to nearest treatment facility | < 15 minutes | 1619 | 29.6 (34.9) | 545 (34) |
|  | 15-30 minutes | 1812 | 32.2 (41.4) | 620 (34) |
|  | > 30 minutes | 839 | 31.5 (38.4) | 272 (32) |

**Table 2:**
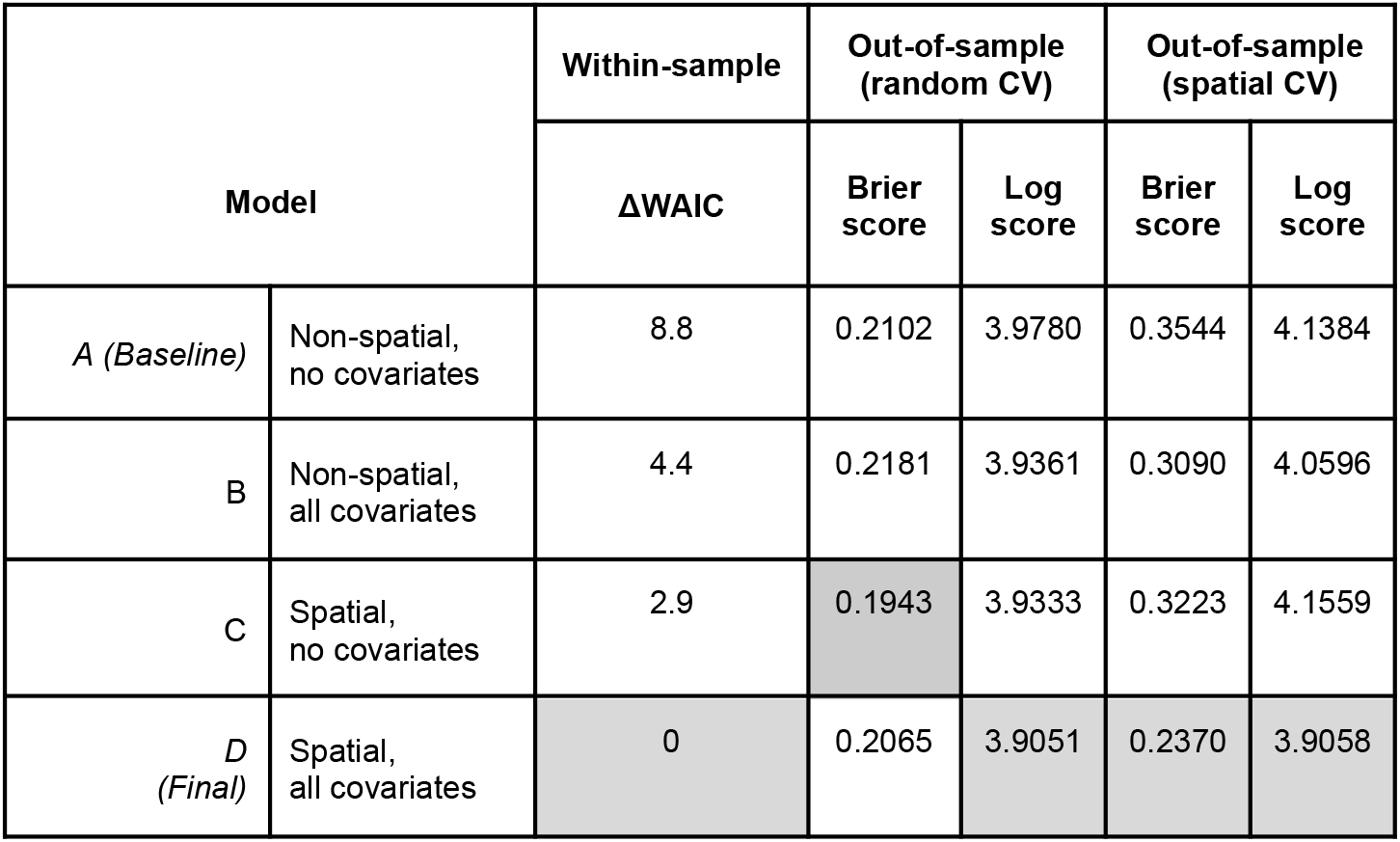
Model comparison on within-sample and out-of-sample fit. The minimum of each metric is shaded in grey. The difference in WAIC (ΔWAIC) between each model value and the minimum is presented as opposed to the absolute value.

### Variable selection

Among patient-specific covariates, age, HIV status and detection by ACD were found to be associated with length of delay (estimated RRs and 95% CrI of 1.14 [1.13,1.15], 1.54 [1.31, 1.81] and 0.74 [0.69, 0.79] in univariate analyses, respectively; *Figure 2*). No clear association was found for caste status or VL/PKDL treatment history, with the direction of effect switching between univariate and multivariate analyses.

**Figure 2:**
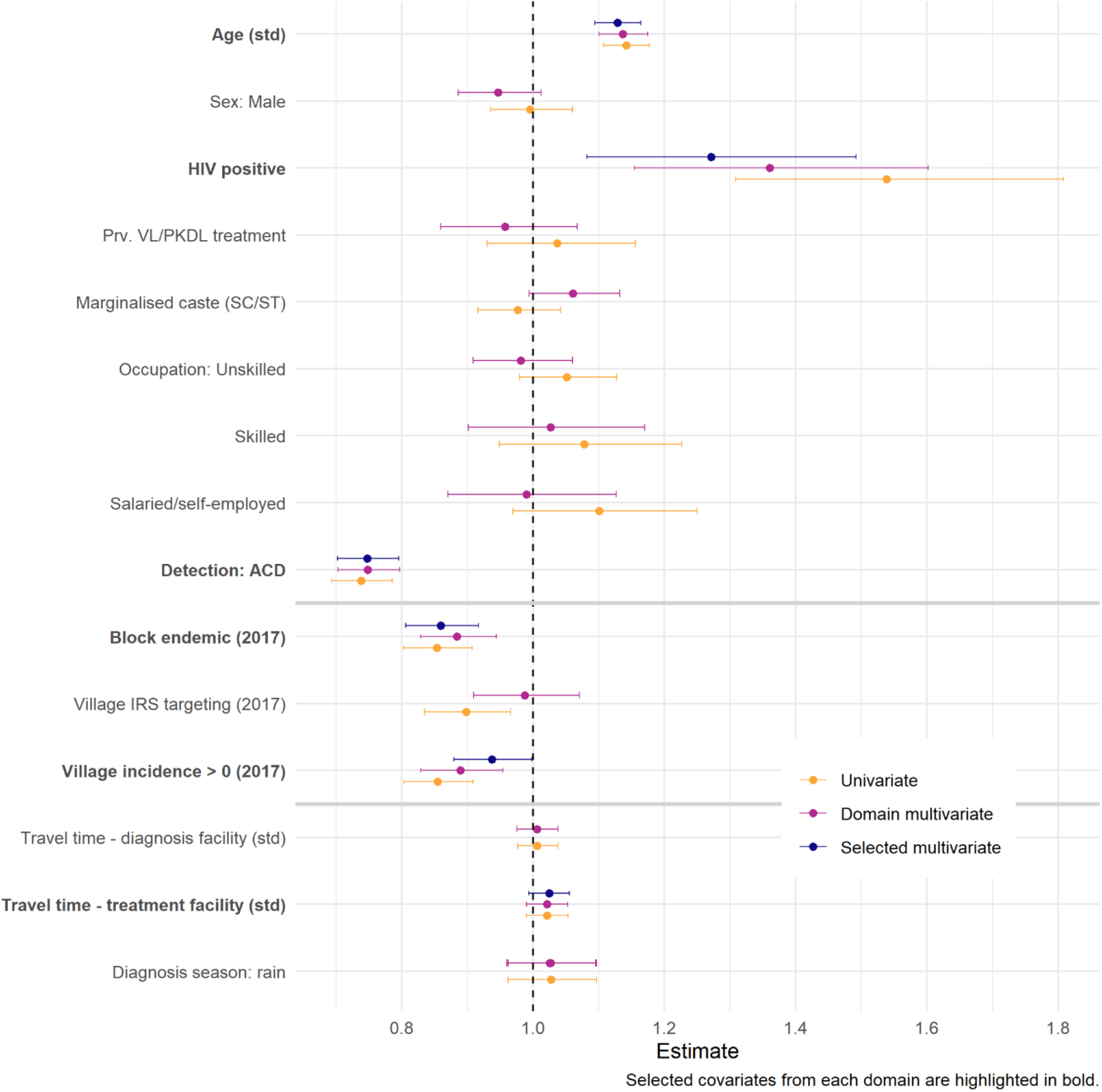
Coefficient estimates (with 95% CrI) obtained from non-spatial model fits: univariate, multivariate within each covariate domain and multivariate with selected covariates from all domains. Selected covariates are also highlighted in bold on the y-axis. Note that domain models were fit to include either travel time to diagnosis or to treatment facility - but not both - due to collinearity in these covariates (closest diagnosis facility may also be closest treatment facility), therefore the domain coefficient for diagnosis season is estimated twice.

Within the “risk awareness” domain, block endemicity and non-zero village incidence in the previous year were associated with shorter delays. Estimated RRs for these two covariates were very similar in univariate analyses (0.85 [0.80, 0.91] and 0.86 [0.80, 0.91], respectively), suggesting that they may capture some of the same variation. Although IRS targeting had a negative effect in univariate analysis, this was lost when accounting for the other covariates in the domain (adjusted RR 0.99 [0.91, 1.07]).

Within the “access” domain, no clear univariate associations were found. When travel time was combined with season in multivariate analyses, time to treatment facility (in minutes) had a borderline positive association with delay (1.02 [0.99, 1.05]). For completeness, this covariate was selected for comparison of all three domains in later analyses.

### Spatial analysis and final model

Incorporating an explicit spatial structure in the residuals alongside the chosen covariates yields the lowest WAIC out of all models compared (*Table 1*). Gains on out of sample prediction are also evident, with respect to both log score and Brier score on predicting exceedance of 30 days.

The estimated covariate effects from the final, spatial model were consistent with those from the non-spatial model (*Supplementary Figure S4*). Being aged 1 SD above the mean of 28 years and being HIV positive were associated with a 13% (95% CrI [9.3% - 16.0%]) and 28% [9.2% - 49.4%] increase in delay, respectively. Diagnosis via active rather than passive case detection was associated with a 22% [17.9% - 26.8%] reduction in delay. In terms of local awareness of VL, patients residing in blocks considered endemic and villages with non-zero incidence in the year prior to diagnosis experienced 13% [2.9% - 21.7%] and 7% [1.3% - 13.1%] shorter delays, respectively, after adjusting for the sources of individual level variation described above. The final model gave some indication of an increase in delay with longer travel time to a treatment facility however the evidence for this remained weak.

The fitted spatial effect had a posterior range (the approximate distance beyond which correlation falls below 0.1) of 47km (95% CrI [26km - 84km]), and a standard deviation of 0.32 [0.23 - 0.42]. The SD of the OLRE decreased from 0.99 [0.97 - 1.01] in the null model to 0.96 [0.94 - 0.99] in the non-spatial model, and finally to 0.93 [0.90 - 0.95] in the final, spatial model, as more of the residual variance could be explained by other components. Converting to a binomial likelihood to compensate for heaping did not substantially alter the inferred relative effects of the covariates (*Supplementary figure S5*).

*Figure 3 (A)* illustrates the spatial pattern of diagnosis delays estimated from the final model, assuming diagnosed cases are comparable on all factors apart from location. *Figure 3 (B)* translates these projections to exceedance probabilities, mapping the estimated probability of observing delay greater than 30 days at any location. Less opaque areas indicate where the probability is close to 0.5 and hence exceedance of 30 days is least certain. The pattern highlights regions in the north west (across Siwan, Gopalganj and Paschim Champaran districts), north east at the Nepal border (Supaul and Araria districts), and further south (Patna, Vaishali and Munger) across which delays are on average expected to be longer than 30 days. It also flags more focal regions of possible concern around Saraiya (Muzaffarpur district), Kalyanpur (Samastipur) and Sonbarsa (Sitamarhi) blocks. This pattern differs from that observed in total incidence (*Figure 3 (C)*); the cluster of higher incidence blocks between the Ghaghara and Gandak rivers northwest of Patna is not reflected by a comparable cluster in the distribution of diagnosis delays.

**Figure 3:**
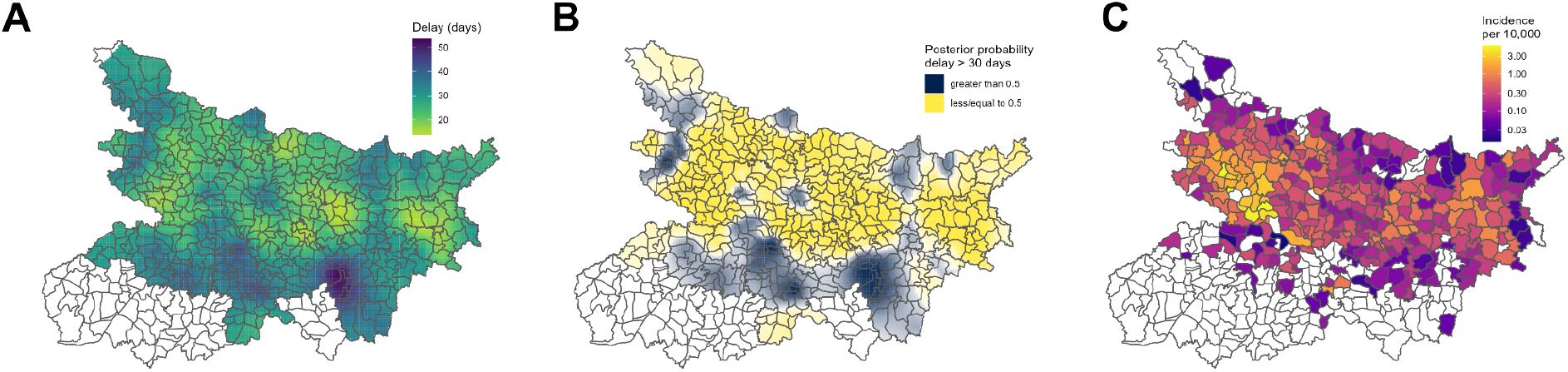
**(A)** Model-estimated spatial variation in delay,assuming that cases are comparable on all factors except location. **(B)** Probability of these predicted delays exceeding 30 days, categorised to highlight where probability is greater than (yellow) or less than (black) 0.5. The opacity of colour reflects distance of the estimate from 0.5 i.e. the strength of the classification. **(C)** Observed total block-level incidence per 10,000. Note: Estimates are not mapped for districts within which no cases were observed during the period of the study.

The map of predicted exceedance probabilities is illustrated in *Supplementary figure S6,* alongside an alternative to Figure 3B using a higher cut-off of 0.75.

### Impact of ACD

In total over all observations, predicted total person-days of delay was reduced by just under 15% when ACD coverage was increased to 100% of cases, equating to a reduction of 7.7 (98% CrI [5.5 - 9.8]) days per case among those originally detected by passive case detection (PCD) (*Figure 4*). This reflects a reduction of 8.7 [6.2 - 11.0] days per reassigned case in non-endemic blocks, compared to only 6.7 [4.8 - 8.6] days in endemic blocks. By increasing ACD detection from its current value of 40.1% to 100%, the overall average estimated delay decreased by 4.6 days, from 31.5 to 26.9. See *Supplementary table S4* for a full table of estimates.

**Figure 4:**
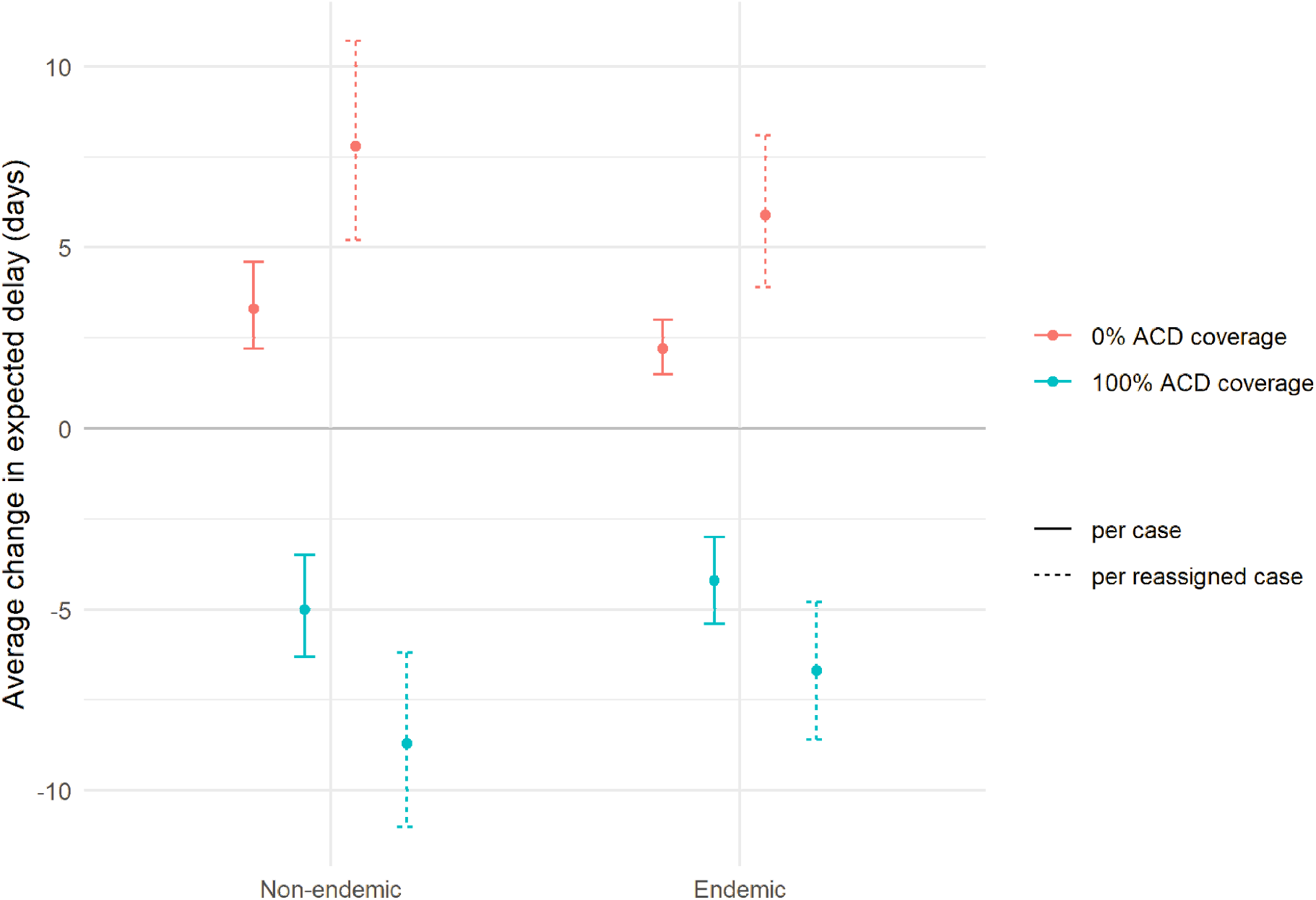
Change in expected diagnosis delay under different ACD coverage scenarios, stratified by recent block endemicity. Baseline is taken as the expected delay under the actual coverage observed in this population. Estimates are shown as average days per case in total and average days per case for which detection route was reassigned under the scenario (i.e. those originally ACD in the 0% scenario, and those originally PCD in the 100% scenario). Point estimates are medians and intervals are 98% credible intervals over 10,000 posterior samples.

Conversely, in the complete absence of ACD (0% of cases), estimated total person-days of delay increased by around 9% - an average difference of 7 [4.6 - 9.6] days per case among those originally detected by ACD. The difference between endemic and non-endemic blocks is also clear in this scenario, with a greater increase observed in endemic blocks (7 [4.6 - 9.6] days per reassigned case) than in non-endemic blocks (6.3 days per reassigned case). In the absence of any ACD, the average estimated delay for all VL cases increased by 2.8 days.

## Discussion

In any disease elimination setting, a new set of challenges arises as incidence is suppressed to very low numbers. The effort required to detect each individual case grows rapidly, yet it is at this stage - when immunity and attention are potentially waning - that prompt detection is crucial to avoid resurgence. Sparsity of incidence across a broad geographic area prompts focussing attention and resources on specific areas considered to be most at risk based on recent observed data. However, this reactive approach may have unintended consequences for the observation of incidence, biasing surveillance because of feedback between case detection and detection effort.

This work highlights a geographic pattern (with a range of around 50km) to the villages in which cases experience the longest delays and motivates further investigation to understand what drives this pattern. Currently, areas of concern are identified for intervention according to recent observed incidence. However, ACD could be more effective if guided not only by incidence but by where delays are longest or most problematic for transmission. In all model fits, ACD was found to be strongly related to the time taken to obtain a diagnosis, associated with greater than 20% shorter delay on average than PCD. We estimated that if all cases in this study had been detected actively, the total person-days of delay accumulated during this period may have been reduced by nearly 15%. This translated to a reduction of 5 days per case in recently endemic blocks versus 4.2 in recently non-endemic, suggesting that gains from active detection in terms of person-days of delay avoided may be greater across recently non-endemic than endemic blocks. Characterising this spatial variation offers guidance to areas in which there is greatest scope to reduce delays - and hence understanding transmission risk - through increased coverage of active surveillance.

The inferred relationship between the length of delay and recent incidence in the region could reflect the impact of waning awareness and detection effort in areas which have not been recently affected. This concurs with previous work investigating variation in seeking of and access to VL diagnosis. A study conducted in 2019 in Nepal compared samples of districts included and excluded from the national control programme and found increased delays in care-seeking among patients in non-programme districts ^20^. Awareness and attitudes around VL have been evaluated in various settings, with one study concluding that this may affect the likelihood of treatment-seeking through appropriate channels ^21^ and another finding understanding to be lacking even among individuals having experienced VL in their household^22^. The possibility should be considered that both the benefit of ACD and promptness of independent care-seeking may wane as we move closer to elimination.

Focusing attention on areas considered “high risk” from recently observed incidence may risk delaying diagnosis and treatment among the few cases which arise in low-endemic areas. This could be a concern since recent evidence has arisen of increasing, sporadic incidence of VL beyond the main endemic regions ^23^. A study in Vaishali district by Kumar and colleagues ^24^ suggested the need to extend active efforts of vector control, case detection and community engagement to non-endemic but high-risk villages peripheral to hotspot areas. However, the authors conceded that there are substantial economic barriers to applying this intensive approach.

ACD is laborious and the cost severely limits its viability in areas with no recently reported cases. Yet, Dial and colleagues ^25^ make the case that bolstering efforts in meso-endemic and low-endemic districts may prove to be cost-effective in the long term. We found evidence that ACD may have greater scope for reducing delay in low-endemic areas. As communities from low-endemic areas lack awareness to promptly recognise symptoms and self-refer, it provides further justification for maintaining robust surveillance in such areas. However, it should be considered whether there is a more economical approach to active surveillance than its current form. A cost assessment could, for example, be made of a strategy not to intensively detect cases but to intensively increase awareness outside the assumed endemic areas (awareness of the disease, its diagnosis/treatment and of PKDL).

If the past decade of efforts continues to be successful and incidence declines to near-negligible levels in many districts, our findings suggest that this may result in longer delays for the few remaining cases. Medley and colleagues ^26^ suggest that prompt diagnosis may be key for India to follow the examples of Nepal and Bangladesh in achieving elimination as a public health problem, but there is scope for further investigation of the consequence of delays among few cases on risk of outbreaks and resurgence. Key epidemiological features ought to be carefully and regularly monitored as programme objectives are achieved, generating feedback with which to periodically update procedures.

Our study has few limitations. Self-reported symptom durations are prone to bias; the raw data exhibit heaping at rounded time intervals and literature suggests that this behaviour can bias parameter estimates ^15^. However, refitting the final model for a binary outcome only reduced the precision of estimates rather than altering the estimated effects. The subset of observations not linkable to GPS locations or with other missing characteristics could also bias the observed spatial pattern of delays or estimated covariate effects. Moreover, the grouping of individual observations by village could mask or dilute important associations. It is the intention of KAMIS data managers that, going forward, each patient’s data would be linked to an individual household location as opposed to only the village centre. The increased identifiability of these data would, however, need to be carefully navigated to take advantage of this finer information for the purposes of surveillance and analysis.

Our interpretation of ACD impact assumes no unobserved confounding in estimation of the intervention’s effect. ACD is triggered by incidence in the last 12 months therefore this is a strong candidate for confounding but is adjusted for with block and village level indicators in the model. A more rigorous analysis of ACD specifically, which considered assumed causal relationships between covariates in more detail, may better pinpoint where and in which populations its benefit might be greatest relative to the cost.

This analysis only describes the behaviour of symptomatic infection among detected cases. The observed delay data may under-represent the upper tail of the distribution since presence in the dataset is conditional on having recognisable symptoms and obtaining a diagnosis at all. The majority of infections with *Leishmania donovani* are asymptomatic and resolve without intervention ^27^, yet xenodiagnostic evidence suggests that asymptomatic individuals do not contribute substantially to transmission ^28^. If poorer detection of symptomatic cases overall corresponds with less prompt diagnosis as observed here, the absence of as-yet-undetected cases from the analysis could render our results conservative and suggest that inferred areas of longer delay could reflect an even greater problem in practice.

Also excluded are cases of PKDL, a more poorly-reported secondary form of leishmania infection which may contribute increasingly to transmission as VL incidence declines ^29^. Delays to diagnosis of PKDL are usually longer than for VL, yet may exhibit similar spatial patterns since detection of PKDL can be a by-product of VL surveillance.

Reduction of avoidable delays to diagnosis and treatment is a key objective in the pursuit of visceral leishmaniasis elimination across the Indian subcontinent. Previous work has identified some groups at risk of delayed care-seeking, but we demonstrate that heterogeneity remains in the promptness of diagnosis across the state. This spatial variation may in part be explained by differences in risk awareness because of recent VL incidence in the community. Evidence suggests that returns on active detection may vary between regions at different stages of elimination, and we suggest that further mathematical modelling may clarify how delays could perpetuate transmission in low incidence areas. The efficacy of active case detection in reducing delays is clear, yet its intensity and geographic extent may need to be reassessed as the region approaches elimination.

## Contributors

ESN, OJB and GFM conceptualised the study. ESN performed the analyses, wrote the manuscript and produced the remote code repository. OJB and GFM provided supervision, advised on the study concept and provided feedback on the manuscript. TL provided feedback on the methodology and presentation of results. JB, PD and KB were responsible for collection, cleaning and maintenance of the data. CB and SS supervised the data collection. MMC supervised as PI of the SPEAK India consortium. All co-authors provided feedback on the manuscript and approved the final submitted version.

## Data sharing statement

The full analysis dataset cannot be publicly shared as it contains both sensitive (HIV infection) and identifiable (age, sex, and GPS of resident village) information on individual patients. An altered dataset with GPS locations jittered in order to not correspond to unique villages can be made available upon request. It should be noted that from this it would not be possible to exactly replicate the presented results.

## Acknowledgments

The authors offer their profound gratitude to the field teams at CARE India who work tirelessly to collect and organise the case details and spatial data without which this work would not have been possible, and the National Centre for Vector Borne Diseases Control (New Delhi, India) for their permission to use the data.

