## Supplementary Materials for "Spatial variation in delayed diagnosis of visceral leishmaniasis in Bihar, India"

#### A Mesh construction

A triangular mesh on which to base the spatial Stochastic Partial Differential Equation (SPDE) model was constructed such that the distance between nodes was between 2km (the average distance between nearest-neighbour affected villages) and 10km.

#### B Cross-validation

Fifty iterations of spatial and non-spatial cross-validation were performed, to assess the contribution of the random field to prediction. For the former, all observations within a 50km radius of a randomly sampled point were withheld from model fitting and delay for the sampled point then predicted. For the latter, only the sampled point was withheld and then predicted. A cross-validated logarithmic score (logs) was calculated across all fifty test observations, summarising the log posterior density at the observed value. Classification of delays greater than 30 days was also assessed via the Brier score, defined as the mean squared difference between the posterior probability of delay exceeding 30 days (the *exceedance probability*) and the observed (binary) value.

### Figures

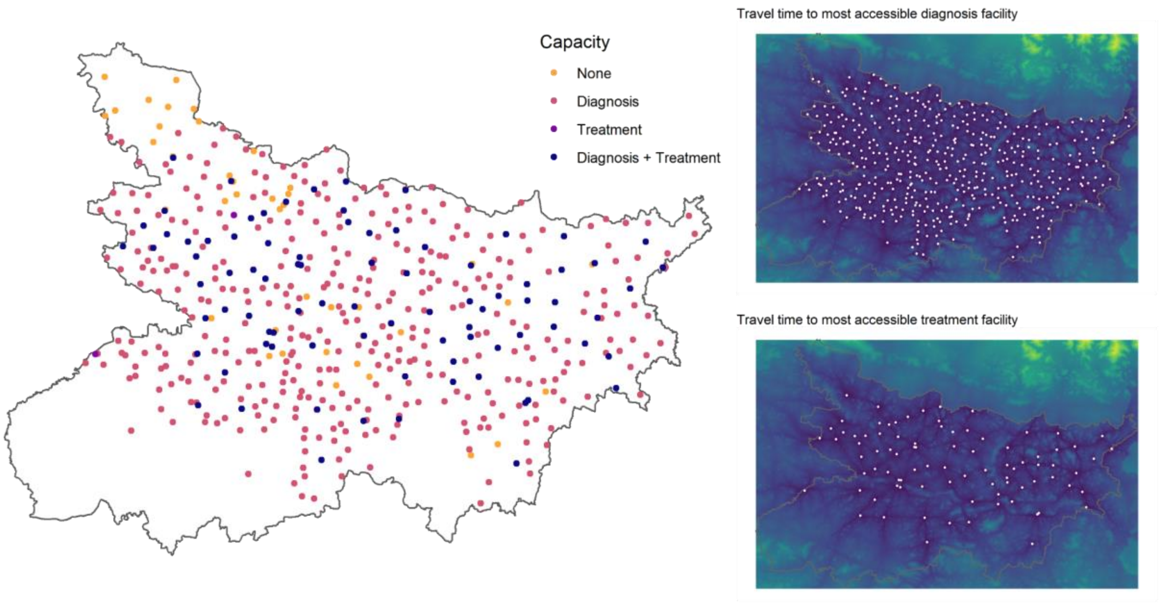


***Supplementary figure S1:*** *Locations of health facilities in 33 endemic districts of Bihar, with capacity for diagnosis and/or treatment of visceral leishmaniasis (VL). Minimum estimated travel time to one of these facilities from any point in the state is illustrated in the panels on the right.*

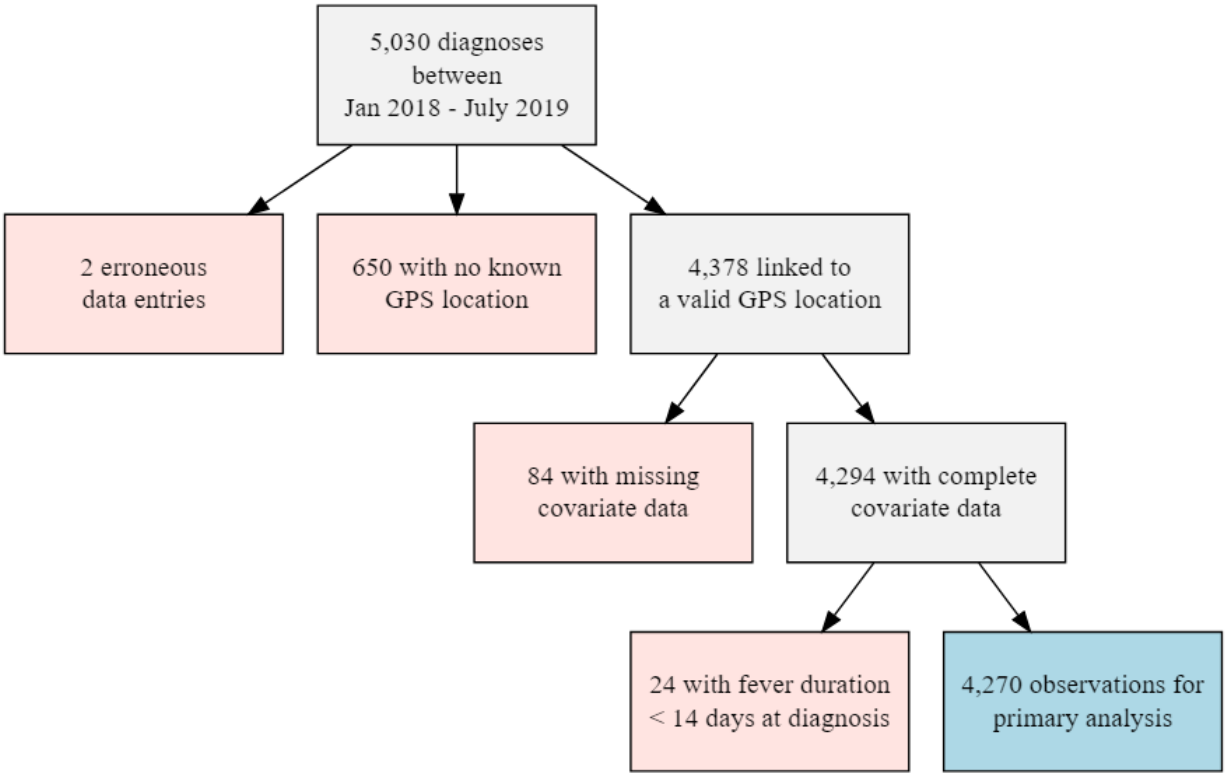


***Supplementary figure S2:*** Flow chart of the data cleaning process, illustrating number of observations excluded under each criterion and the remaining observations included in the primary analysis.

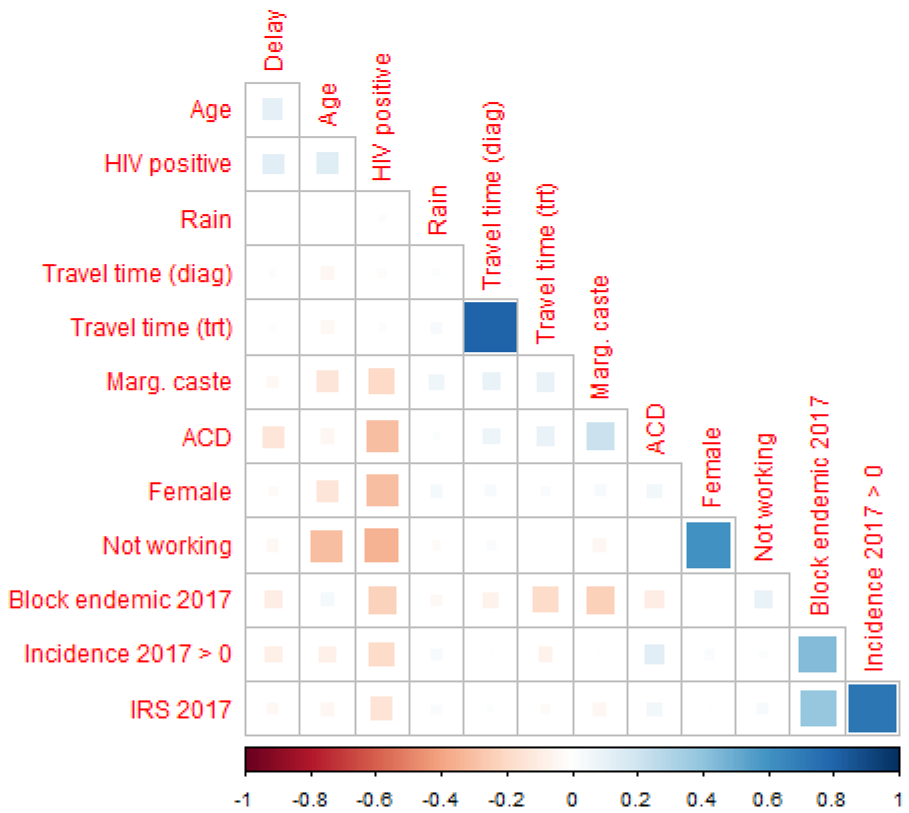


***Supplementary figure S3:*** *Correlations between diagnosis delay and all covariates considered. The point-biserial method is used for correlation between binary and continuous variables, and the Pearson method otherwise. The size of the square for each combination indicates the strength of correlation, while the colour indicates both strength and direction. The strongest correlations are observed between travel time to diagnosis facility and travel time to treatment facility, as to be expected since several facilities provide both services, between village vector control and incidence in 2017, and between employment and sex.*

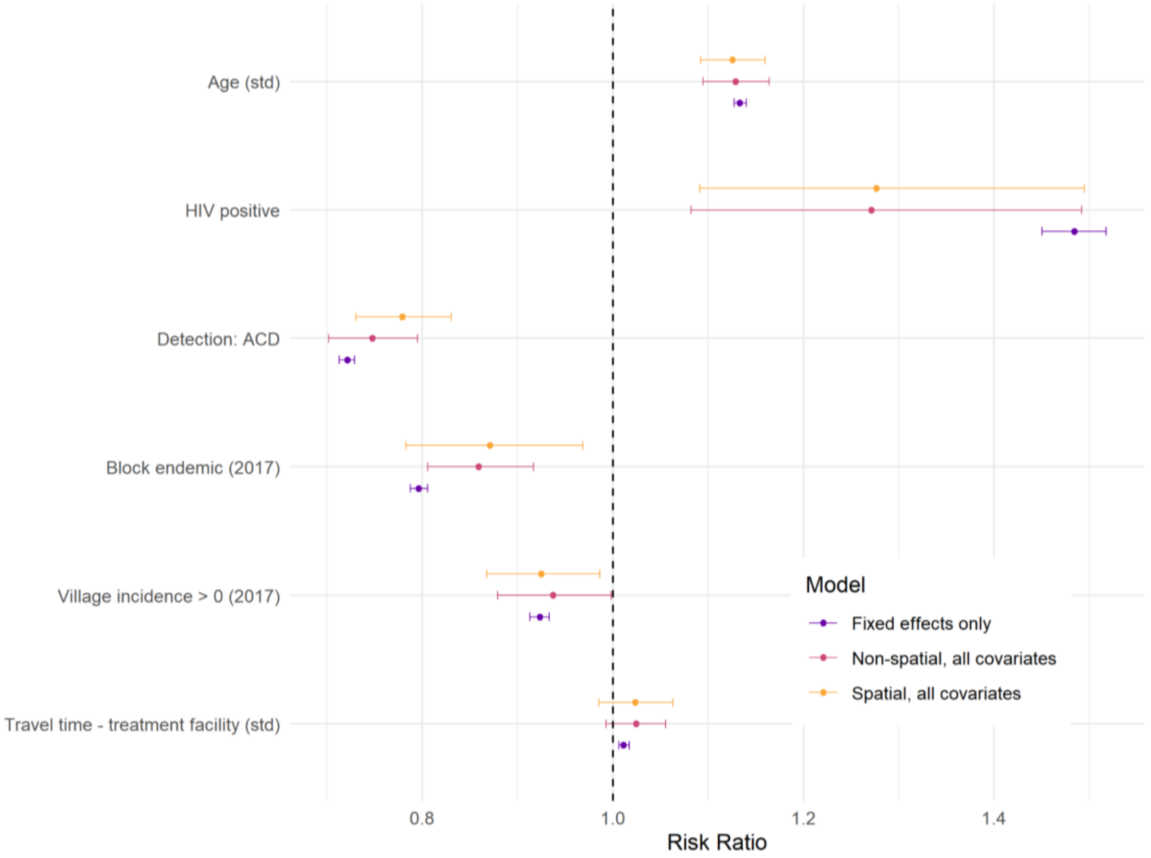


***Supplemental figure S4:*** *Comparison of coefficient estimates between non-spatial and spatial models. Estimates are also shown from a model with only fixed effects. All significant covariates remain so with addition of the spatially-structured random effect. The estimated effect size for detection route and historical block endemicity are marginally reduced, while that of historical village incidence is increased. ACD - active case detection.*

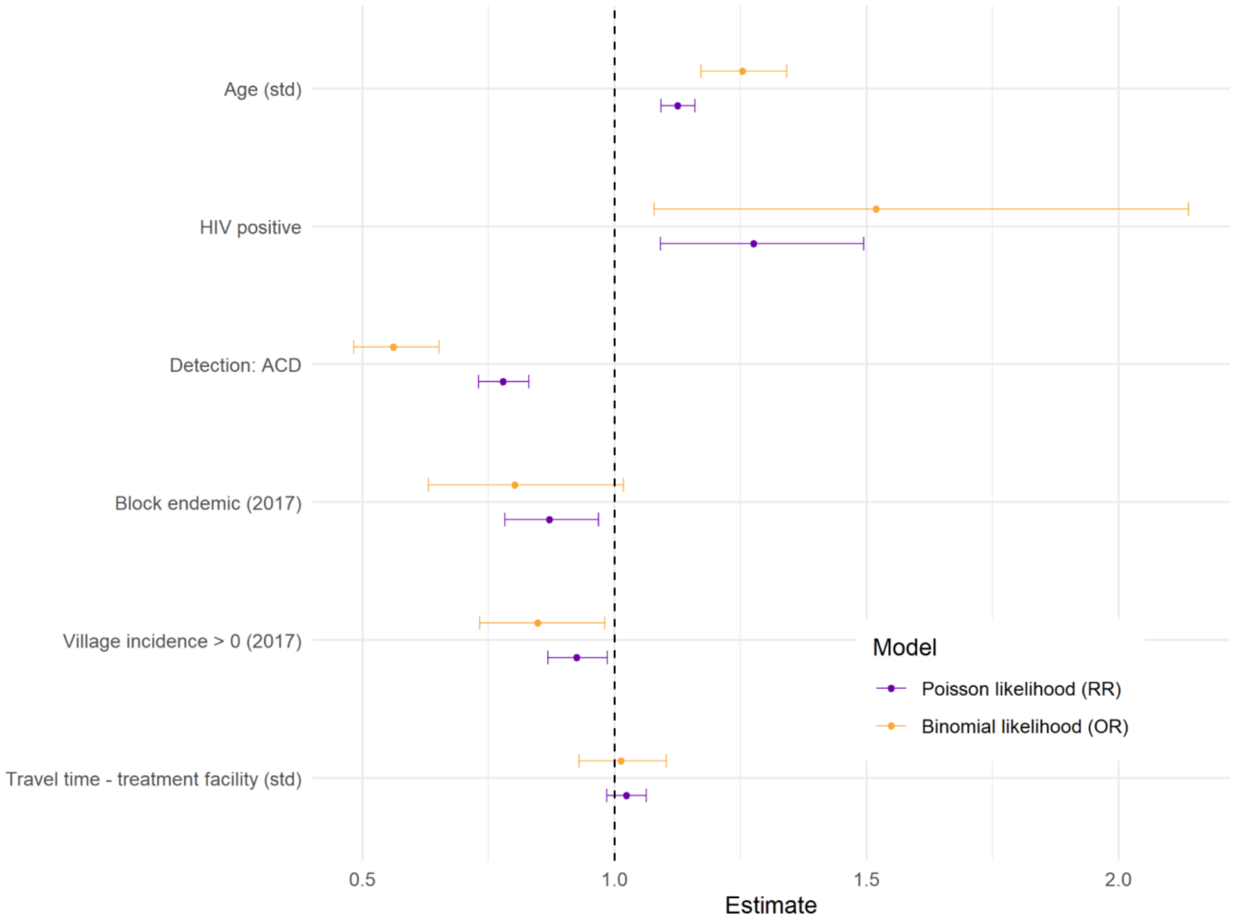


***Supplementary figure S5:*** *Comparison of coefficient estimates (with 95% Credible Intervals) between final model (Poisson likelihood) and an alternative using a binomial likelihood with a cut-off of 30 days' delay. It is important to note that estimates from the final model represent risk ratios while for the alternative binomial model they represent odds ratios. Therefore the magnitude of effects cannot be directly compared, only the direction.*
*RR - risk ratio; OR - odds ratio.*

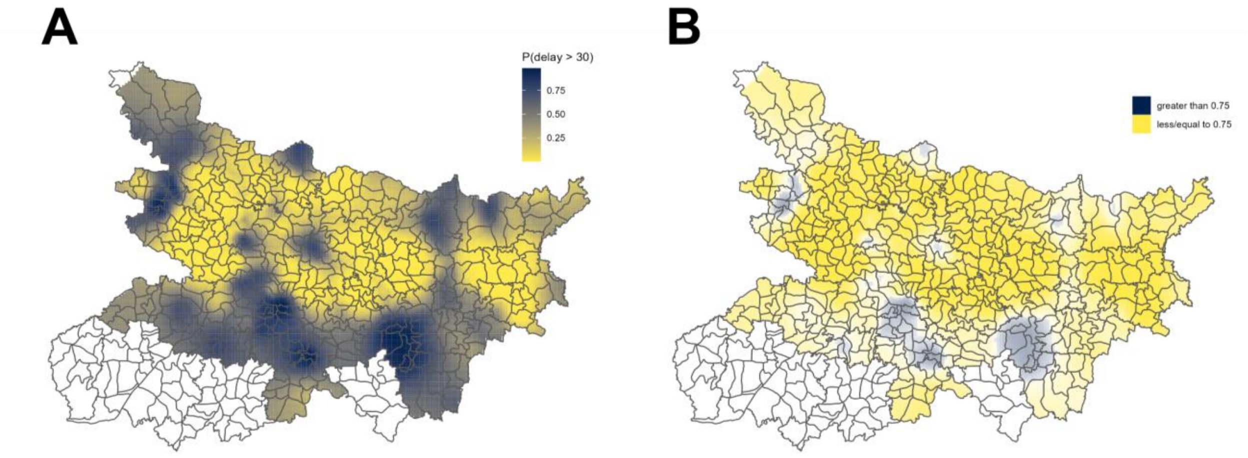


***Supplementary figure S6: (A)*** *Predicted probabilities of delay exceeding 30 days and* ***(B)*** *an alternative version of Figure 3B with a higher cut-off of 0.75, to illustrate the impact of the choice of cut-off value. Delays greater than 30 days are only expected with high probability in limited regions of the south/south-east and northwest.*

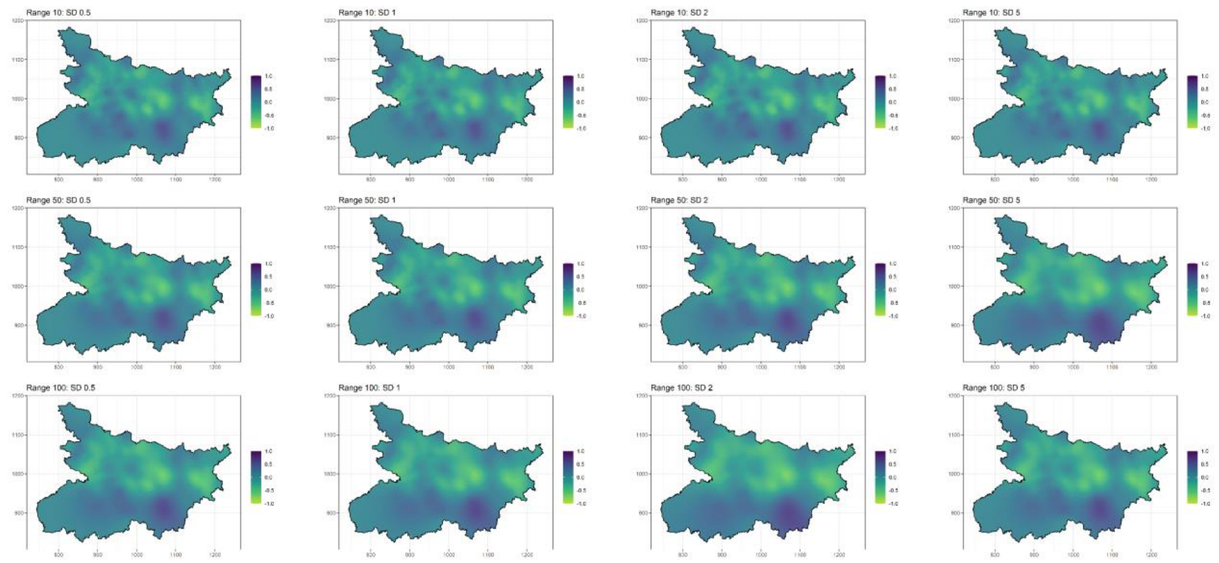


***Supplementary figure S7:*** *Sensitivity analysis of the SPDE prior specification, comparing fitted spatial fields from the final model, with varying prior range and standard deviation.*

### Tables

***Supplementary table S1:*** *Summary of 736 observations out of 5,030 which were excluded from the dataset prior to analysis, due to either missingness of GPS location for the village or missingness in one or more of the individual-level covariates of interest.*

| | **Excluded** | **Included** |
|---|---|---|

| | | |
|---|---|---|
| N | 736 | 4294 |
| Delay, median [IQR] | 16 [11-46] | 16 [11-44] |
| Age, median [IQR] | 28 [13-45] | 25 [12-42] |
| Female, n (%) | 285 (38.7) | 1833 (42.7) |
| HIV positive, n (%) | 241 (33.3) | 1498 (34.9) |
| HIV missing, n (%) | 12 (1.6) | 0 (0) |
| Previous VL/PKDL treatment, n (%) | 35 (5.2) | 158 (3.7) |
| Previous treatment missing, n (%) | 69 (9.4) | 0 (0) |
| Marginalised caste, n (%) | 59 (8.1) | 377 (8.8) |
| Caste status missing, n (%) | 8 (1.1) | 0 (0) |
| Unemployed, n (%) | 399 (55) | 2525 (58.8) |
| Missing occupation, n (%) | 11 (1.5) | 0 (0) |
| Diagnosed through ACD, n (%) | 277 (37.6) | 1720 (40.1) |

VL - visceral leishmaniasis; PKDL - post-kala azar dermal leishmaniasis.

***Supplementary table S2:*** *Summary of patient and village level characteristics of cases reporting less than or greater than 14 days of fever prior to diagnosis. Intervals presented are approximate 95% confidence intervals for mean or percentage.*

| | **Duration of fever before diagnosis** | |
|---|---|---|
| | **< 14 days** | **>= 14 days** |
| N | 24 | 4270 |
| Age | 16 [9-30] | 25 [12-42] |
| Female, n(%) | 4 (16.7) | 1829 (42.8) |
| Marginalised caste, n(%) | 6 (25) | 1492 (34.9) |
| HIV positive, n (%) | 0 (0) | 158 (3.7) |
| Previous VL/PKDL treatment, n (%) | 3 (12.5) | 374 (8.8) |
| Unemployed, n (%) | 19 (79.2) | 2506 (58.7) |
| Diagnosed through ACD, n (%) | 7 (29.2) | 1713 (40.1) |
| Resident of village with non-zero VL incidence in 2017, n (%) | 12 (50) | 2341 (54.8) |
| Resident of village targeted for IRS in 2017, n (%) | 16 (66.7) | 3277 (76.7) |
| Resident of village in an block classed as endemic in 2017, n (%) | 17 (70.8) | 1938 (45.4) |

| | | |
|---|---|---|
| Travel time to diagnostic facility, median [IQR] | 9 [5-12] | 12 [7-18] |
| Travel time to treatment facility, median [IQR] | 16 [11-19] | 18 [11-27] |

IRS - indoor residual spraying

***Supplementary table S3:*** *Changes in magnitude of fitted random effects (non-spatial and spatial) with inclusion of each covariate domain.*

| Included domain | Non-spatial effect (OLRE) | | Spatial effect (SPDE) | |
|---|---|---|---|---|
| | Mean absolute value | % change | Mean absolute value | % change |
| None (*Model C*) | 0.7005 | - | 0.2517 | - |
| Patient (age, HIV, detection) | 0.6889 | -1.66 | 0.2374 | -5.69 |
| Awareness (block endemicity, village incidence) | 0.6978 | -0.39 | 0.2289 | -9.06 |
| Access (travel time to treatment facility) | 0.7005 | 0.00 | 0.2495 | -0.87 |

***Supplementary table S4:*** *Summary of delays associated with active active case detection (ACD) and passive case detection (PCD), by recent block endemicity. The relative gains from ACD compared to PCD appear greater in blocks which had not been recently classified as endemic.*

| | **Non-endemic 2017** | **Endemic 2017** |
|---|---|---|
| No. blocks | 290 | 44 |
| Total population | 76484757 | 9389809 |
| No. cases detected | 2332 | 1938 |
| *via ACD (%)* | 995 (42.7) | 718 (37.0) |
| Mean delay - overall | 34.4 | 27.0 |
| *via ACD* | 27.8 | 20.8 |
| *via PCD* | 39.5 | 30.7 |
| Difference PCD-ACD (Std. err.) | 11.7 (1.8) | 9.8 (1.4) |

***Supplementary table S5:*** *Estimated total person-days of delay (median and 98% credible interval over 10,000 posterior samples) in the scenario of (a) complete ACD coverage, i.e. with all observations redefined as actively detected and (b) no ACD coverage, i.e. with all observations redefined as passively detected. The impact of the detection scenario is summarised with respect to the change in total person-*

*days of delay relative to the original fitted values, as an overall total and split by the endemicity of the block.*

| Scenario | Measure | Total | Non-endemic 2017 | Endemic 2017 |
|---|---|---|---|---|
| Observed ACD coverage *(Baseline)* | No. blocks | 334 | 290 | 44 |
| | No. detected cases | 4270 | 2332 | 1938 |
| | Via ACD (%) | 1713 (40.1) | 995 (42.7) | 718 (37.0) |
| | Expected total person-days delay | 134 631 [133 760, 135 495] | 81 345 [80 675, 82 000] | 53 283 [52 739, 53 829] |
| | Per case *(ACD + PCD)* | 31.5 [31.3, 31.7] | 34.9 [34.6, 35.2] | 27.5 [27.2, 27.8] |
| Modelled complete (100%) ACD coverage | Expected total person-days delay - | 114 793 [109 454, 120 650] | 69 772 [66 603, 73 230] | 45 042 [42 779, 47 478] |
| | Per case *(originally ACD + PCD)* | 26.9 [25.6, 28.3] | 29.9 [28.6, 31.4] | 23.2 [22.1, 24.5] |
| | Change from baseline | -19 811 [-25 180, -14 118] | -11 575 [-14 693, -8 256] | -8 234 [-10 479, -5 851] |
| | Per case *(originally ACD + PCD)* | -4.6 [-5.9, -3.3] | -5 [-6.3, -3.5] | -4.2 [-5.4, -3] |
| | Per reassigned case *(originally PCD only)* | -7.7 [-9.8, -5.5] | -8.7 [-11, -6.2] | -6.7 [-8.6, -4.8] |
| Modelled no (0%) ACD coverage | Expected total person-days delay | 146 645 [142 519, 151 121] | 89 114 [86 421, 92 047] | 57 530 [55 974, 59 195] |
| | Per case *(originally ACD + PCD)* | 34.3 [33.4, 35.4] | 38.2 [37.1, 39.5] | 29.7 [28.9, 30.5] |
| | Change from baseline | 12 009 [7 942, 16 437] | 7 761 [5 129, 10 612] | 4 246 [2 812, 5 813] |
| | Per case *(originally ACD + PCD)* | 2.8 [1.9, 3.8] | 3.3 [2.2, 4.6] | 2.2 [1.5, 3] |
| | Per reassigned case *(originally ACD only)* | 7 [4.6, 9.6] | 7.8 [5.2, 10.7] | 5.9 [3.9, 8.1] |
